# Computational Methods for Characterizing Research Outputs, Collaborative Networks and Thematic Concentration: a Case Study in Primary Care Research Evaluation

**DOI:** 10.1101/2023.09.07.23295220

**Authors:** Christopher Meaney, Peter Selby, Mary Ann O’Brien, Ross Upshur, Jaya de Rege, Rahim Moineddin, Yuxi Lily Ren, Selena Ma

## Abstract

**Objective:** Research impact is difficult to measure, evaluate and report. This study aims to demonstrate how computational scientometric methods, including bibliometric, network analytic, and thematic summary measures can efficiently characterize complex scientific disciplines, such as primary care research.

**Methods:** We used a retrospective cohort design. The study included N=17 international academic primary care research departments. A scientometric database was curated using a bottom-up methodology, which included peer-reviewed research articles/reviews, and associated meta-data, published between 01/01/2017 and 31/12/2022. Publication-level bibliometric information was queried from the Scopus application programming interface (API). The Altmetrics API was used to extract publication-level indicators of social engagement. Network analytic visualizations and statistics characterized research collaboration. Topic models and keyword mining characterized the main thematic areas of primary care research. At an author-level, we investigated correlations between bibliometric, altmetric, network analytic and topical summary measures.

**Results:** Our analysis included N=591 primary care researchers (from 17 institutions) who produced 13,047 unique peer-reviewed articles over the study timeframe. These 13,047 research articles were published in 2,237 unique journal titles; cited 231,121 times; and received broad social uptake (605,349 Twitter tweets, 36,982 mainstream media mentions, 884 Wikipedia references, and 1,127 policy document citations). The 591 researchers collaborated with 35,585 unique co-authors resulting in 20,808,886 pair-wise collaborations. The median number of authors per publication was 7 (IQR: 4-10; min=1; max=3,391). Frequently occurring keywords/n-grams and latent topical vectors, highlighted the diversity of primary care research. Clinical research themes included: physical/mental health conditions, disease prevention and screening, issues in primary/obstetric/emergency/palliative-care, and public health. Methodological research themes included: research synthesis/appraisal, statistical/epidemiological inference, study design, qualitative research, mixed methods, health economics, medical education, and quality improvement. Many themes were stable over the study timeframe. COVID-19 emerged as an important research theme from 2020 through 2022. Topic vectors encoding clinical medicine were positively correlated with bibliometric, altmetric and network centrality measures, whereas, vectors encoding qualitative methods, medical education, and public health were negatively correlated with these same metrics.

**Conclusions:** Multi-metric, computational scientometric methods offer an efficient, transparent, and reproducible means for characterizing the research output of complex scientific disciplines, such as primary care research.

## 1 Introduction

At the individual-, organizational-, and system-levels stakeholders are interested in collectively assessing research outputs/impacts and generally the returns associated with research investments. Aggregate research assessments can be operationally challenging to design and implement. The assessments can be time-consuming and costly to conduct, oftentimes involving subject-matter experts with specialized skills in research evaluation. Current best practices suggest a balanced approach to research evaluation/assessment, involving a mix of quantitative performance indicators in addition to complementary qualitative analyses (e.g. narrative/oral summaries obtained from stakeholder surveys, interviews, or focus groups) [Thomas et al., 2020, Cabezas-Clavijo and Torres-Salinas, 2021]. This study used computational methods to curate and analyze aggregate scientometric data, providing a multi-metric lens for the purpose of research program evaluation, assessment and benchmarking. We illustrate the relative ease by which bibliometric, altmetric and related information can be mined using popular data scientific computing languages (e.g. R or Python). The general and adaptable methodology we illustrate can be used to characterize any complex research discipline; however, in this study we focus on the application of a computational scientometric approach for the evaluation, assessment and benchmarking of primary care research.

Primary care is a foundational component of healthcare systems across the globe. Primary care aims to maintain/improve patient health through the provision of a diverse range of promotive, protective, preventive, curative, rehabilitative, and palliative health services [AAFP, 2023]. Primary care is multi-disciplinary with a focus on the biological, behavioural, and social determinants of patient health/wellbeing. Research is a mechanism to foster the evidence-informed development of primary care. Ideally, research ensures that the best science is translated into primary care practice, fostering construction of a learning primary healthcare system. Primary care research program leadership, primary care research funders, and primary care researchers themselves, are interested in the characterization of their research discipline. At an individual-level, primary care research characterization can guide informed participation in targeted research activities, whereas, at a system-level, it can inform coordination, investment, and other capacity building activities. Indicators of scientific output (i.e. scientometrics) represent a general methodological framework for measuring and characterizing primary care research. Metrics of scientific output/performance are readily used in the context of academic program evaluation and decision making [Abbott et al., 2010, Thomas et al., 2020]; and there exist several examples of their use in primary care research as well. For example, Glanville et al. [2011] and Liaw et al. [2019] used bibliometric techniques to summarize primary care research outputs in the United Kingdom and United States, respectively; Hong et al. [2016] used methods from text mining and natural language processing to identify themes of primary care research; and Vezyridis and Timmons [2016] combined methods from bibliometrics, network science and keyword analysis to characterize primary care research in the United Kingdom. In this study, we seek to investigate how a computational scientometric methodology, involving several complementary data sources and statistical methods, can be used to characterize recent primary care research activities.

The overarching goal of this research study is to characterize current primary care research conducted at several international primary care research institutions using a multi-metric scientometric methodology [Martin, 1996, Cabezas-Clavijo and Torres-Salinas, 2021, Szomszor et al., 2021]. Particularly, we illustrate how modern computational technologies can be leveraged to efficiently, transparently, and reproducibly curate a primary care publication database (consisting of publication data, and associated bibliometric and altmetric meta-data). And further we demonstrate how these large/diverse scientometric data can be married with modern statistical methods, to generate unique insights for stakeholders interested in primary care research assessment/evaluation.

## 2 Methods

### 2.1 Study Design, Setting and Selected Institutions/Authors

Our illustrative case study in primary care scientometrics used a retrospective cohort design. We used a “bottom-up” sampling framework [Van Leeuwen, 2007]. We started by sampling individual researchers, and collecting all of their associated scientific research articles/reviews. Inferences regarding higher level research units (e.g. researchers or institutions) were generated by aggregating over individual article/review data associated with a particular stratification of interest. Through a consensus building process, several members of the research leadership team from the University of Toronto Department of Family and Community Medicine identified N=17 high-performing academic primary care research programs for inclusion in our scientometric study (see Table 1). Representation included academic primary care research institutions from the following countries: Canada (N=5), the United Stated of America (N=6), the United Kingdom (N=3), Australia (N=2), and Hong Kong (N=1). Primary care researchers were identified from the N=17 research program webpages (see Table 1). Two authors (SM, JdR) independently and manually extracted researcher-name lists from institutional webpages (in the spring/summer of 2022). Two authors (SM, JdR) manually queried Scopus IDs and added valid unique persistent identifiers to the researcher-name lists. Two additional authors (YLR and CM) reviewed the researcher-name lists extracted by SM and JdR, and adjudicated inconsistently extracted names/IDs: resulting in N=658 included primary care researchers. Table 1 summarizes characteristics of the N=17 selected academic primary care research institutions included in our study.

**Table 1:**
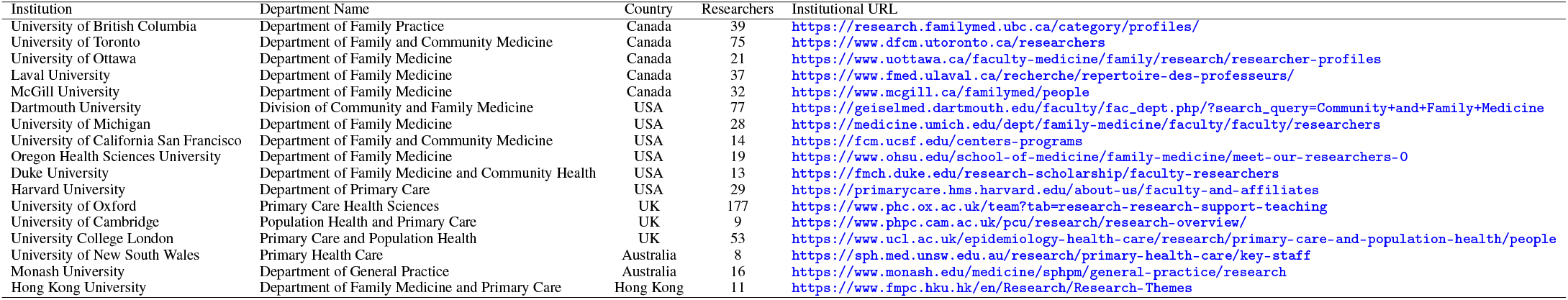
Institutions selected for inclusion in our scientometric study, and a description of their country, research staff/faculty size, and associated research program webpage/URL.

We included researchers generating at least one original research article/review (indexed in the Scopus database) between January 1, 2017 through December 31, 2022 (a 6 year study timeframe). In total, 591/658 (89.8%) researchers published at least one research article/review over our study timeframe, and were included in the final analytic sample.

### 2.2 Data Sources, Data Structures and Curation of a Scientometric Database

The scientometric study involved computationally mining data from two primary sources: 1) the Scopus Application Programming Interfaces (API) from the Elsevier Developer Portal [https://dev.elsevier.com/sc_apis.html], and 2) the Altmetrics API for researchers [https://www.altmetric.com/products/altmetric-api/].

Using the curated list of first/last names collected for each selected primary care research institution, we identified associated Scopus IDs for each author using the following web search interface [https://www.scopus.com/freelookup/form/author.uri]. The Scopus ID is a unique persistent identifier (automatically generated by Scopus) at the researcher-level. A majority of primary care research staff/faculty who have history of producing academic knowledge products have an associated ScopusID. The unique persistent identifiers circumvent downstream issues associated with author name disambiguation.

Using the list of Scopus IDs for each primary care researcher, we adopted a “bottom-up” sampling strategy for bibliometric database curation. The information contained in the database was validated by the team and properly reflect scientific performance [Van Leeuwen, 2007, Zuccala et al., 2010]. For this approach, we engaged with the Scopus API using the unique Scopus IDs and extracted all indexed research articles/reviews generated by each primary care researcher (and associated publication meta-data). We verified the data resulting in deleting certain IDs that were selected erroneously and adding IDs that were previously missed in the data collection, while accounting for duplication. In this study we queried the following information from the Scopus API (using the rscopus package: https://github.com/muschellij2/rscopus):

- Document level ID variables (Digital Object Identifier (DOI), Scopus EID)
- Document Title
- Document Authors
- Document Author Affiliations
- Document Abstract
- Document Keywords
- Document Citation Count

To investigate the social impact of primary care research articles/reviews identified above, we queried the Altmetric API using document-level identifiers (e.g. the DOI) collected from the initial Scopus API call (using the rAltmetric package: https://github.com/ropensci/rAltmetric). The Altmetrics database collects information on a variety of alternative metrics of research quality/impact [Bornmann, 2013, 2015]; we focused on specific Altmetric measures specified below:

- Number of Tweets
- Number of Wikipedia citations
- Number of mainstream media coverage events (e.g. television/radio/newspaper coverage)
- Number of citations in policy documents

### 2.3 Metrics, Statistical Analysis and Reporting

Our scientometric study sought to characterize research outputs, social impact, network collaboration, and thematic concentration of primary care researchers over our study timeframe. Our study inferences were largely descriptive. We adopted a multi-metric evaluation framework utilizing a wide range of bibliometric indicators to assess research outputs and academic impact [Martin, 1996].

To characterize research outputs and academic impact of research, we reported publication and citation counts. This is further complemented with Altmetrics to expand our view and capture new forms of academic impact as expressions of scholarship become more diverse and digital. To capture social impact of research, we reported on various Altmetric indicators including: number of Twitter tweets, counts of mainstream media coverage events, Wikipedia references, and numbers of citations in policy documents.

Using publication co-authorship lists, we constructed a network graph (adjacency matrix). We used network centrality indices to characterize vertex (i.e. author/researcher) importance in the network. In this study we investigate degree centrality, closeness centrality, betweenness centrality and page rank centrality [Newman, 2004, 2010]. We extracted author affiliations (i.e. institution/country) and characterized research studies according to collaboration (local, national, or international).

Using publication keywords we descriptively characterized common primary care research thematic areas. Further, we mined title/abstract n-grams to investigate thematic research areas. We present the top-25 most frequently occurring keywords and n-grams. We constructed a document-term-matrix (DTM) from the publication abstract data, and used non-negative matrix factorization (NMF) to extract a topical basis describing the primary care research corpus [Paatero and Tapper, 1994, Lee and Seung, 2000]. NMF hyper-parameters were optimized via random search (using N=200 experimental configurations), to identify a hyper-parameter configuration which maximized topical coherence [Bergstra and Bengio, 2012, Röder et al., 2015]. We aggregated topical prevalence vectors by 1) publication year, and 2) Scopus Author ID. We explored temporal variation in topical prevalence over our study period. Further, we investigated Spearman correlations between topical/thematic vectors (aggregated at an author-level) and various bibliometric, altmetric and network thematic indicators (also aggregated at an author-level).

### 2.4 Reproducible Research/Program Evaluation

The research name/ID list (including information on N=658 primary care researchers from N=17 academic primary care research institutions); along with all R scripts (Jupyter Notebook files) required to query data from APIs, clean/transform data structures, perform statistical analyses and generate tables/figures, are published at the author’s GitHub repository for this study [https://github.com/meaneych/PrimaryCareScientometrics].

### 2.5 Ethics

This scientometric study was a component of a larger internal research program evaluation exercise conducted at the University of Toronto, Department of Family and Community Medicine. The project was deemed a quality improvement study by the University of Toronto Research Ethics Board, and hence did not require a formal review or research ethics ID assignment.

## 3 Results

### 3.1 Descriptive Statistics of the Primary Care Research Corpus

A total of N=17 academic primary care research institutions, and N=658 primary care researchers, were included in our bottom-up computational scientometric study. N=591 (89.8%) published at least one primary care research article/review (indexed in Scopus) during our study timeframe (01/01/2017 through 31/12/2022). These researchers produced 13,047 unique research articles, which were published in 2,237 unique journal titles. Popular journal titles included: focused primary care journals (e.g. Annals of Family Medicine, Canadian Family Physician, and British Journal of General Practice, etc.), general medical journals (e.g. British Medical Journal, Canadian Medical Association Journal, etc.), and focused medical specialty journals (e.g. Canadian Journal of Emergency Medicine, Palliative Care, etc.). Publication frequency in particular journal titles tended to be associated with geographic location of authors (i.e. authors were more likely to publish scholarly works in journal titles associated with their home countries).

### 3.2 A Topical/Thematic Characterization of the Primary Care Corpus

We present the top-25 most frequently occurring publication keywords, publication title bigrams/trigrams, and publication abstract bigrams/trigrams as a mechanism of characterizing the diversity of primary care research (Table 2).

**Table 2:**
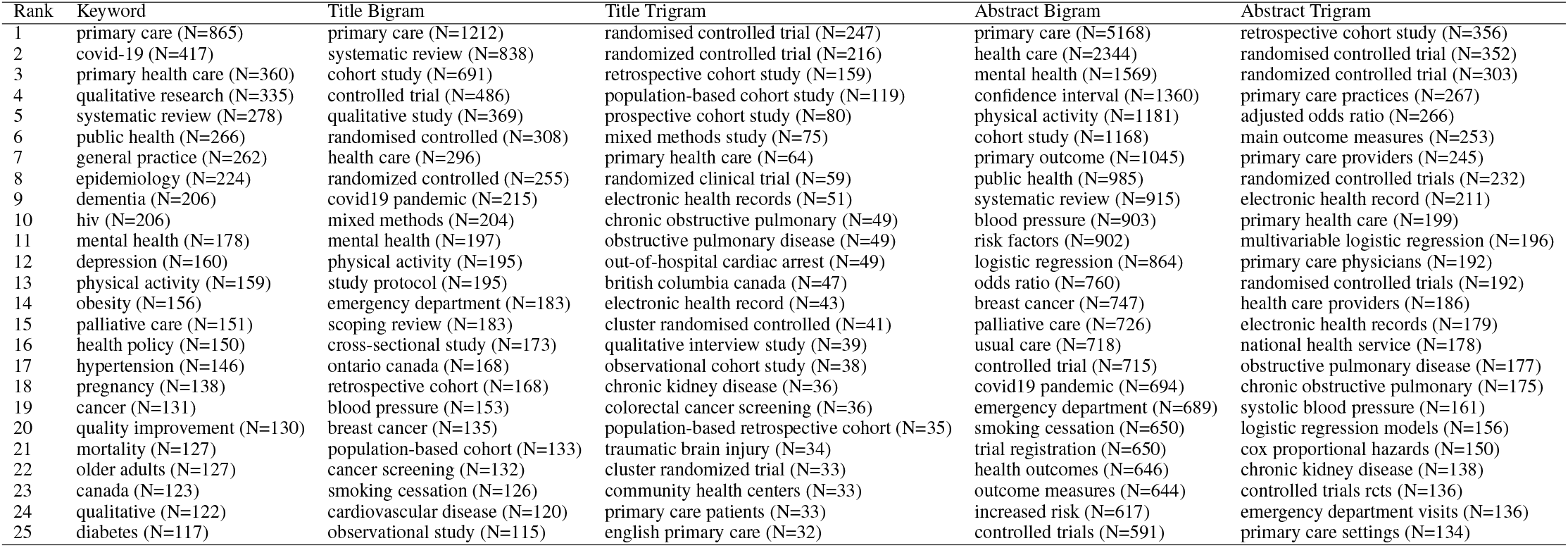
Top-25 most frequently occurring keywords, publication title bigrams/trigrams, and publication abstract bigrams/trigrams.

We fit a NMF topic model (with K=60 topics) to the primary care publication abstract corpus. The analysis further corroborates the diversity of primary care research. A range of identified research themes included: physical/mental health conditions, disease prevention and screening, issues in primary/obstetric/emergent/palliative-care, public health, medical education, and quality improvement. Methodological research themes included: research synthesis/appraisal, statistical/epidemiological inference, principles of study design, qualitative research, mixed methods and health economics. We stratified topical prevalence estimates by study-year and depict the evolution of primary care research topics over our study timeframe. Most topics appear stable over the study timeframe; with a major exception being the clear emergence of a COVID-19 topic becoming of interest in 2020-2022 (see Topic 50 in Figure 1, below).

**Figure 1:**
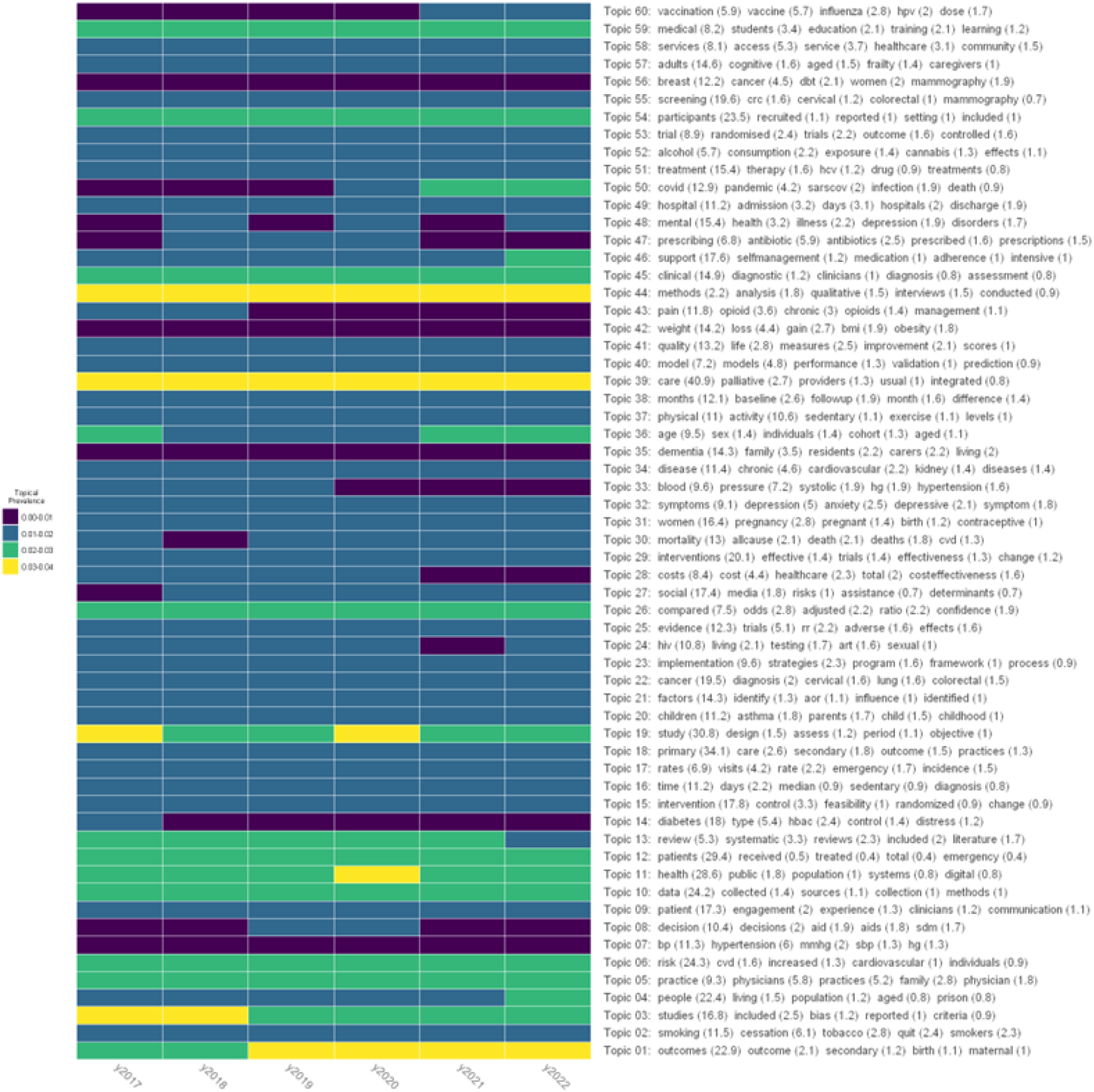
Topical prevalence estimates stratified by study year (2017-2022), estimated from a non-negative matrix factorization topic model with K=60 latent dimensions. Row vectors of the heatmap describe the evolution of a specific topic over our study timeframe; the column vectors describe a topical probability distribution estimated over a single study year.

### 3.3 Characterizing Primary Care Research Collaboration

The N=591 researchers published 13,047 unique research articles/reviews that involved unique 35,585 unique co-authors (resulting in 20,808,886 pair-wise collaborations/co-authorships). The median number of authors per publication was 7 (IQR: 4-10; min=1; max=3,391). 34% of collaborations were international (involving authors from different countries), 43% were national (involving only authors from the same country) and 23% were local (including only authors from the same city). Figure 2 illustrates the highly complex primary care research co-authorship network structure. We computed network centrality measures at an author-level (particularly, degree centrality, closeness centrality, betweenness centrality and page rank centrality) as a mechanism for identifying important individuals within this social network (these are discussed further in upcoming results sub-section).

**Figure 2:**
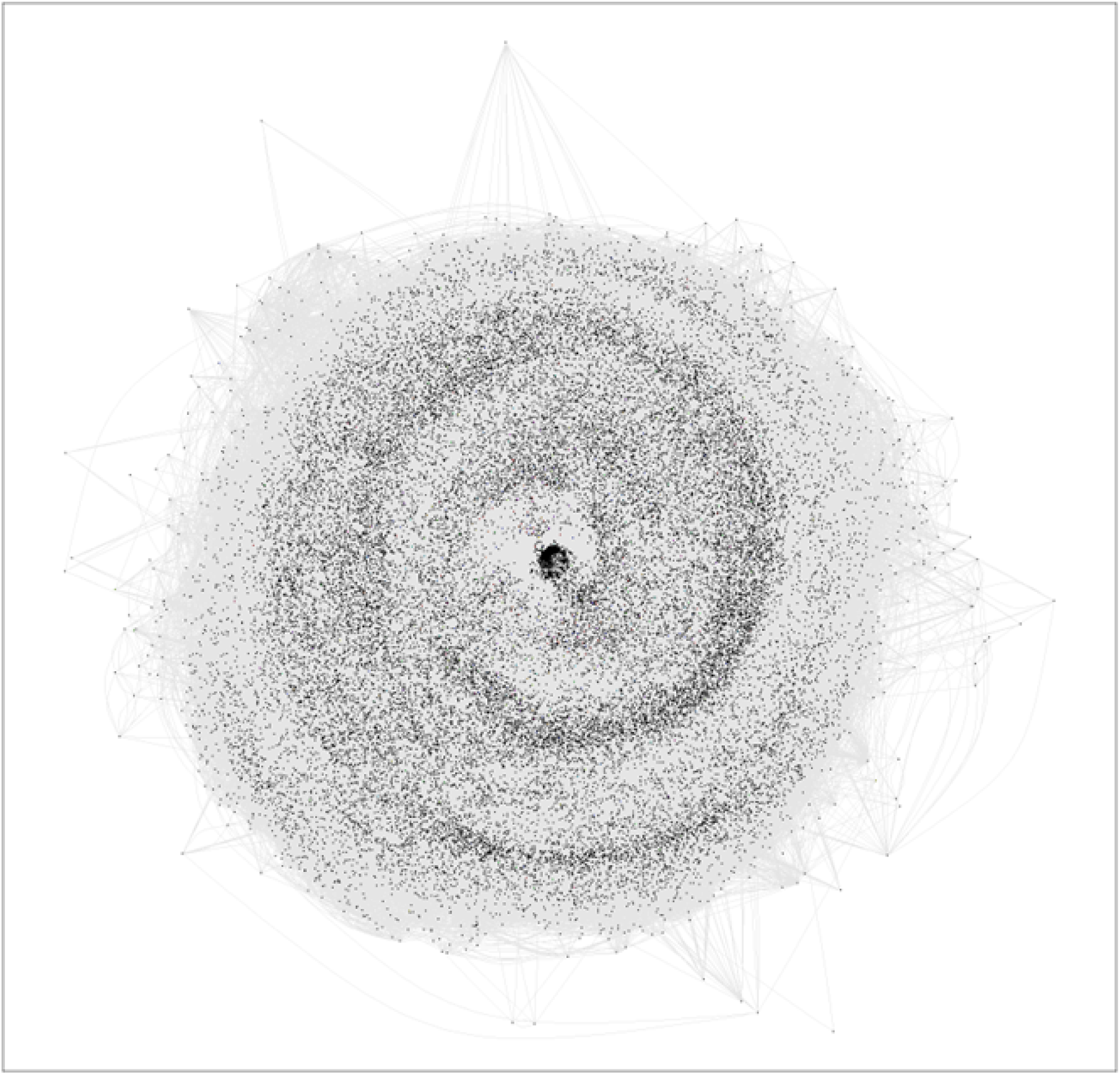
Network diagram illustrating collaborations/co-authorships between primary care researchers involved in this scientometric study. The network diagram includes 36,176 unique nodes/authors (591 primary care authors queried, plus 35,585 unique collaborators). The network consists of 20,808,886 edges representing pair-wise collaborations/co-authorships between researchers.

### 3.4 Bibliometric and Altmetric Indicators of Research Output and Social Impact

The N=591 primary care researchers produced 13,047 unique peer-reviewed articles/reviews over the study timeframe. These 13,047 research articles/reviews (published in 2,237 unique journal titles) were cited 231,121 times. The collection of primary care research received broad social uptake (605,349 Twitter tweets, 36,982 mainstream media mentions, 884 Wikipedia references, and 1,127 policy document citations).

### 3.5 Correlation of Bibliometric, Altmetric and Network Analytic Indicators (on a researcher-level)

We observed that all bibliometric, altmetric and network scientific indicators of research output, social engagement and research collaboration are positively correlated (Figure 3).

**Figure 3:**
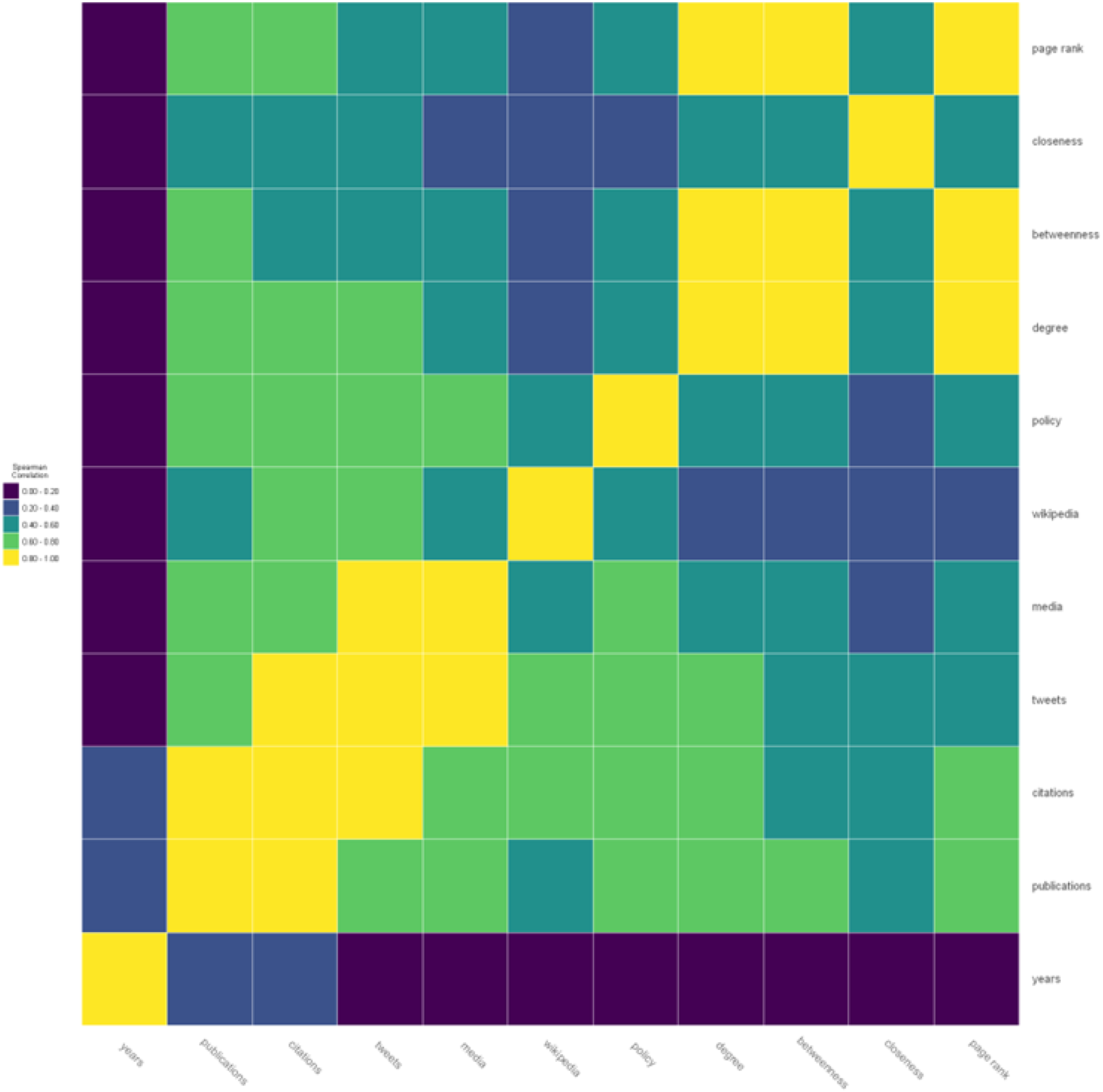
A heatmap illustrating Spearman correlation coefficients between research experience (years), bibliometric, altmetric and network scientific indicators.

We further correlate topical prevalence vectors (aggregated at an author-level) with the aforementioned bibliometric, altmetric, and network scientific indicators. With respect to citations (a measure of research impact) and altmetrics (a measure of social impact) we observe certain interesting positive/negative correlations (Figure 4). For example, topical vectors encoding themes such as qualitative research methods, medical education, medical decision/communication aids demonstrated negative correlations with bibliometric/altmetric indicators. Conversely, topical vectors encoding areas of clinical medicine such as COVID-19, cardiovascular disease and hypertension, asthma, cognition/frailty, and smoking cessation demonstrated positive correlation with bibliometric/altmetric indicators.

**Figure 4:**
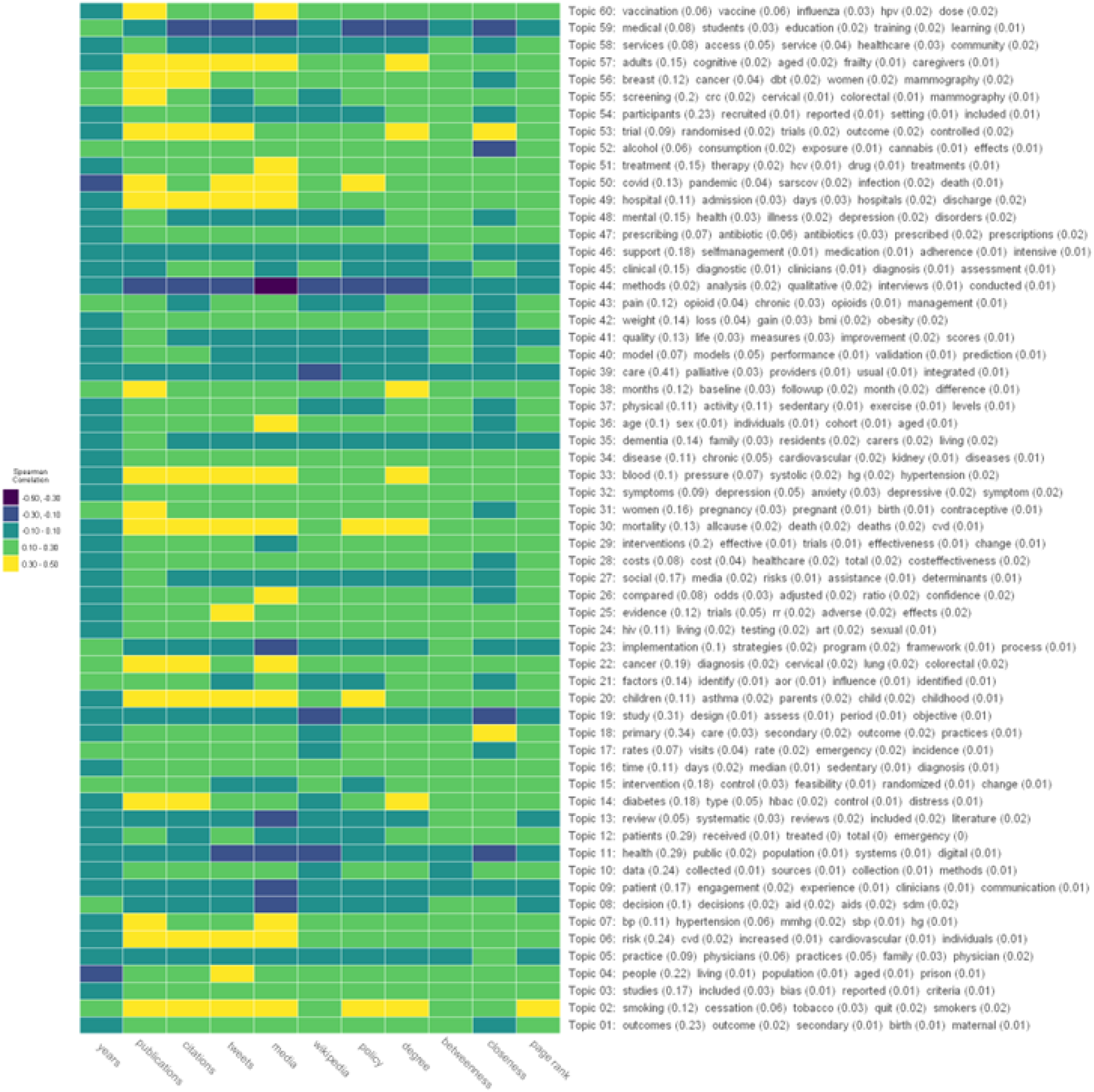
A heatmap illustrating Spearman correlation coefficients between bibliometric, altmetric and network scientific indicators with learned topical prevalence vectors (at an author-level).

## 4 Discussion

This study demonstrated how a computational scientometric methodology can be used to characterize complex scientific disciplines, such as primary care research. The study integrated methods from bibliometrics/altmetrics research, network science, natural language processing, unsupervised machine learning and multivariate statistics. Using a multi-metric approach, we summarized research outputs (publications), academic/social impacts (academic and altmetric citations), networks of collaboration, and major thematic areas of primary care research (and the inter-relationships/correlation between these indicators). The computational scientometric approach was complementary to alternative methodologies for research program evaluation (e.g. narrative reports, qualitative interviews, focus groups and/or surveys).

Mining narrative bibliometric data (e.g. titles, keywords, and research abstracts) allowed rapid characterization of primary care research foci. Clinical research themes included: specific physical/mental health conditions, disease prevention and screening, issues in primary/obstetric/emergent/palliative-care, public health, medical education, and quality improvement. Methodological research themes included: research synthesis, statistical/epidemiological inference, principles of study design, qualitative research, mixed methods, and health economics. Many of the identified research themes were stable over the study timeframe. Temporal topic modelling clearly identified the emergence of an important COVID-19 thematic research area, which emerged in 2020 in response to the COVID-19 pandemic (a finding which has face validity). Further, bibliometric/altmetric indicators support the scientific and social impact of much of the COVID-19 research conducted by primary care researchers over the past three years.

Collaboration in primary care research was strong. In total, the N=591 researchers included in the study published 13,047 unique research articles/reviews that involved unique 35,585 unique co-authors (resulting in 20,808,886 pair-wise collaborations/co-authorships). 34% of collaborations were international (involving authors from different countries), 43% were national (involving only authors from the same country) and 23% were local (including only authors from the same city). The study identified centrally important researchers participating in the primary care research co-authorship network. Further, we investigated correlations between network centrality indices and other bibliometric, altmetric, and thematic summary measures.

The study used traditional bibliometric indicators (e.g. publication and citation counts) as well as altmetric indicators (e.g. Twitter tweets, mainstream media mentions, Wikipedia references, and policy document citations) to characterize academic and social impact of primary care research. We found trends towards increasing research outputs/impacts over time, suggesting the growth and strength of primary care research [AAFP, 2023]. Overall, the academic and social impact of modern primary care research is impressive, with thousands of academic citations and millions of references across various social media and online information platforms.

A novel methodological aspect of this study involved the investigation of correlations between bibliometric, altmetric, network scientific, and topical/thematic summary measures (at an author-level). For example, we identified that all bibliometric, altmetric and network scientific indices displayed positive inter-correlations. This finding is consistent with the work of Eysenbach et al. [2011] who noted a correlation between social impact and academic impact, and Mullins et al. [2020] who reported a significant association between altmetrics and citation count in general surgery. However, such findings are inconsistent [Shiah et al., 2020, Costas et al., 2015, Bornmann, 2015] and difficult to generalize to other specialties [Nocera et al., 2019, Shiah et al., 2020, Kolahi et al., 2021]. Sud and Thelwall [2014] also reported factors, such as journal prestige, type of altmetric, and novelty of the article content as factors influencing metric correlation. To assess academic impact, altmetrics should be viewed as complementary to traditional bibliometrics. To our knowledge, this is the first study to investigate topic models (aggregating topical prevalence vectors from a bottom-up approach at an author-level) and investigate their correlation with the aforementioned bibliometric, altmetric and network scientific indicators. The analysis revealed authors with research foci which were more/less correlated with indicators of research output, research impact and research collaboration. The analysis revealed that topical vectors encoding clinical medicine were positively correlated with bibliometric, altmetric and network centrality measures; whereas, topical vectors encoding qualitative methods, medical education, and public health illustrated negative correlations with the above metrics [Loder et al., 2016]. This may indicate a bias towards quantitative research traditions and support arguments that qualitative research is under represented in medical journals.

A strength of the study was its computational multi-metric approach, which allowed for an efficient characterization of modern primary care research. The methodology allowed for the summarization of research outputs/impacts, collaborative networks, and topical/thematic foci. The computational approach is scalable, auditable, reproducible and transparent. Further, the analytic scripts can be easily modified and extended to permit rapid analysis and programmatic evaluation of other research disciplines (i.e. the methodology can be generalized beyond a characterization of primary care research). Future work should continue to investigate how modern statistical techniques can be applied to curated scientometric databases (including bibliometric, altmetric, journal metric, etc. information) to yield unique insights about researchers, research institutions and/or research disciplines.

### 4.1 Limitations

Our computational scientometric analysis of primary care research is not without limitations. Below we highlight some of the limitations of our study.

From a methodological perspective, we attempted to marry a number of complementary statistical methods to provide holistic insights regarding a research discipline, in particular primary care research. We have not been exhaustive in including/representing all possible statistical techniques which can be applied to scientometric databases in this study. Future best practices should attempt to identify specific statistical methods which can be applied to different data sources/structures which can be routinely extracted from modern scientometric databases.

From a data/measurement perspective, we have included specific scientometric data sources in this study. In terms of data sources, we opted to use the Scopus API to extract bibliometric outputs and the Altmetrics API to collect social engagement metrics. There exists a rich landscape of data curators who allow user engagement with scientometric data for research purposes [Cabezas-Clavijo and Torres-Salinas, 2021]. Ultimately, data accessibility, coupled with documentation and ease of use of the APIs, influenced decisions regarding data curation pipelines. Other data sources may result in slightly different inferences regarding primary care research. Considering bibliometric information, competitor data sources include PubMed, Web of Science, Google Scholar and others. Considering Altmetrics, there also exist PlumX metrics and the Dimensions API. Comparisons of different Altmetric data sources are discussed in Zahedi and Costas [2018] and Ortega [2018]. Further research should continue to investigate variation in study inferences as a function of input scientometric data sources.

From a study design perspective, certain decisions may have impacted inferences regarding our characterizations of modern primary care research. Most importantly, our study only involved researchers from N=17 academic primary care research institutions (N=591 researchers). This is not a complete representation of all primary care researchers (many primary care researchers have been excluded from our analysis; as we could not extract all researchers from all primary care research institutions). If other stakeholders were interested in contributing primary care research data to this analysis, the method would scale easily (however, would require curating additional Scopus IDs on researchers from other institutions). Further, the sample is purposefully biased to include research institutions producing voluminous quantities of primary care research (so is not representative of primary care research conducted in all academic institutions globally).

It is possible that a “bottom-up” vs “top-down” methodology may result in subtly different inferences. This study employed a “bottom-up” methodology for research program evaluation, in that primary units (researchers were identified), and researcher-level information was queried, which was then aggregated up to an institution-level to permit comparative inferences. Alternatively, there exist bibliometric tools to facilitate more “top-down” analyses based on unique persistent research institution IDs, or conducting queries which limit to specific institutions, geographic locations, etc. In the primary care scientometric literature review we have seen examples of both: Glanville et al. [2011] used a “top-down” approach, whereas, Liaw et al. [2019], Van Leeuwen [2007], Zuccala et al. [2010] and others used “bottom-up” approaches.

Finally, we have only reported on a subset of potentially relevant scientometric, bibliometric, altmetric, and network scientific measures. We purposefully showcased a multi-metric evaluation [Martin, 1996, Cabezas-Clavijo and Torres-Salinas, 2021, Szomszor et al., 2021]. We have included bibliometric, Altmetric, network analytic and thematic summarizations in our primary care scientometric investigation. The purposeful use of multiple metrics was to generate many diverse lenses from which to characterize the primary care research landscape. We further wish to advocate for the responsible use of the metrics generated in this report [Wilsdon et al., 2015]. Research and research programs are more than simple metrics. And as advised, we have attempted to embed these metrics-based evaluations into a more balanced mixed methods evaluation of our primary care research program (not relying on a purely computational scientometric evaluation/assessment; despite certain advantages associated with the methodology).

## 5 Conclusions

Holistic, multi-metric, computational scientometric methods offer an efficient, transparent, and reproducible means for characterizing the research outputs of complex scientific disciplines, such as primary care. Future work should showcase how modern computational tools, coupled with complementary statistical techniques can be used to facilitate research program evaluation (e.g. comparative benchmarking of institutions or research disciplines).

## Data Availability

All data produced are available online at: https://github.com/meaneych/PrimaryCareScientometrics

